# Barriers and facilitators of Highly Active Antiretroviral Therapy (HAART) adherence among HIV-positive Women in Southern Ethiopia: A Qualitative study

**DOI:** 10.1101/2024.06.12.24308289

**Authors:** Alemayehu Abebe Demissie, Elsie Janse van Rensburg

## Abstract

**Background:** Adherence to Highly Active Antiretroviral Therapy (HAART) medication is the major predictor of HIV/AIDS treatment success. Poor adherence to HAART creates the risk of transmitting HIV, deteriorating health conditions, treatment failure, increased occurrences of drug-resistant HIV, morbidity and mortality. The objective of this study was to explore and describe the experiences of HAART adherence among HIV-positive women in Southern Ethiopia.

**Methods:** Semi-structured in-depth interviews (IDIs) were conducted with 12 HIV-positive women in Southern Ethiopia who are adhering and non-adhering to HAART. Interviews were conducted in the local Amharic language and audio recorded with permission from the participants. The interviews were transcribed verbatim, coded for themes, categories and sub-categories and analyzed using a thematic data analysis technic.

**Results:** The findings of the study reflected two themes: barriers and facilitators of HAART medication adherence among HIV-positive women. Barriers and facilitators were further categorized into 5 categories. These included patient-related factors, treatment-related factors, psychosocial-related factors, family and community-related factors, and healthcare services-related factors. These categories were further divided into 22 sub-categories. Busy schedule, forgetting the doses, rituals of religion, economic constraints, drug side-effects, pills burden and size, misconceptions about HIV, negative attitudinal disposition towards HAART, refusal to adhere to HAART, depression, lack of hope and courage, stigma and discrimination, relationship with healthcare providers, a working day of HAART clinic, and long waiting time were identified as barriers to HAART adherence. While, family responsibilities, reminder devices, dosage formulation, perceived benefit of HAART, family support, adherence to supporting peer groups, and adherence to counselling/education were identified as facilitators of HAART adherence.

**Conclusions:** Adherence to HAART medication is a major challenge among HIV-positive women in Southern Ethiopia. Therefore, tailored strategies to enhance HAART medication adherence should be targeted addressing the barriers identified in the study.

## Introduction

The accessibility of antiretroviral therapy (HAART) for HIV/AIDS patients has increased rapidly globally, reaching 28.7 million in 2021 from 7.8 million in 2010. However, only 68% of these individuals are virally suppressed, indicating sub-optimal HAART adherence [1]. As less viral suppression is associated with sub-optimal HAART adherence, the global less viral suppression (only 68%) in HIV-positive individuals indicates that sub-optimal (poor) adherence to HAART is a global challenge in HIV/AIDS treatment [2]. Ethiopia, one of the most affected countries by HIV epidemics, has a high prevalence of HIV among women, with 61.1% of all HIV-positive individuals being female [3]. In 2017, there were 414,854 patients, including 21,146 children under 15, taking HAART and the national ART need was 551,630 for adults in Ethiopia[4]. Good adherence to HAART is necessary for achieving the best virological response and reducing drug resistance risk, morbidity and mortality [5, 6]. A research study conducted in Ethiopia with 339 people diagnosed with HIV revealed that 25.4% had poor adherence to their medication [7]. Similarly, another study done in Ethiopia showed that only 73.1% of patients were adherent to their HAART [8]. Various factors contribute to poor adherence, including patients and their families, socioeconomic factors, medication, and health providers and systems. Studies have shown that women face numerous challenges in ensuring adherence to HAART, such as education, age, income, stigmatization, and socio-familial support [9]. Women are also responsible for household activities, which can further disadvantage them in balancing family and social life [10]. There is a lack of studies conducted on HIV-positive women’s adherence to HAART in Ethiopia specifically. To address this issue, programs and care providers need to tailor a combination of feasible interventions to maximize adherence of women to HAART based on individual barriers and opportunities globally and in Ethiopia. Therefore, to address the gap in treatment adherence, this study aims to develop a better insight into adherence among HIV-positive women on ARV treatment in Southern Ethiopia.

## Methodology

### Design and setting

The study used an exploratory qualitative research design to gain new insights and understandings based on the themes, categories and sub-categories that emerge from the data.

The study applied the consolidated criteria for reporting qualitative research (COREQ), which is a set of criteria for reporting qualitative research [11].

### Study setting and sampling procedures

The researchers used a purposive and convenient sampling procedure to select the Southern Ethiopia region and the three urban hospitals that were studied. This sampling technique was chosen to select the southern region from the nine regions of Ethiopia and the study hospitals from 21 urban public hospitals found in Southern Ethiopia based on convenience (most accessible to and easily reached by the researchers) and HIV-positive individuals’ load. The purpose of the selection of these three urban public hospitals was due to their high volume (load) of HIV-positive individuals who were taking HAART as compared to other health facilities in Southern Ethiopia

Women in Southern Ethiopia who are HIV positive and adhering or non-adhering to HAART were purposively and conveniently sampled based on the eligibility criteria. Twelve participants (six adhering and six non-adhering women diagnosed with HIV) were purposively and conveniently sampled for the in-depth interviews. Two more interviews were performed to ensure data saturation was reached.

### Interview tool development and data collection procedure

A semi-structured interview guide was developed based on the literature reviewed. An in-depth interview was chosen as a data collection method for this study because the research topic is sensitive, and it would enhance the confidentiality and anonymity of participants. The intended data were collected from women in Southern Ethiopia who are HIV-positive and adhering or non-adhering to HAART. The in-depth interviews were regulated by the interview guide, and probing questions were asked where more detail was needed. Field notes allowed the researchers to remember the behaviours, impressions, nonverbal actions, and environmental contexts that would not be adequately captured via the audio-tape during the in-depth interviews.

### Data collection and analysis

We conducted the in-depth interviews with 12 HIV-positive women. The IDIs were audio-recorded (with the participant’s permission). The interviews were conducted in Amharic, a local language most people in Ethiopia speak. The in-depth-interviews last approximately 60-70 minutes. The researchers used field notes in the in-depth interviews to substantiate the findings from this qualitative data-gathering technique [12]. Data were collected from June 3 to 25, 2023 until data saturation was reached. The researchers translated the audio-recorded data from Amharic into English for analysis. Data from in-depth interviews and field notes were analyzed using the thematic analysis method, which is a search across a data set (interviews, field notes, or a range of data) to find repeated patterns of meaning. The researchers used the Nvivo 12 qualitative data analysis computer program for data analysis [13]. The researchers used thematic analysis for the current study since it is recommended as a convenient method for employing within a participatory research paradigm with participants as collaborators [14]. Finally, some verbatim quotes from HIV-positive women are used to illustrate the sub-categories.

### Ethical Considerations, Rigour, and Trustworthiness

This study obtained scientific approval from the scientific committee (Department of Health Studies and ethical clearance (Ref: HSHDC/463/2015) from the College Research Ethics Committee of the University of South Africa (UNISA). Similarly, permission to carry out the study was asked for and gained from the stakeholders, including the Southern Regional Health Bureau and the three public hospitals where the study was conducted. Confidentiality was ensured at all stages of the process. Informed consent was obtained from the participants prior to the interviews. Participants gave written informed consent before enrollment using the approved consent forms. Participants were assured that refusal to participate or withdrawal from the study would not affect their access to healthcare services at the clinic. The researchers considered the essential values of ethical research, which incorporate respect for human autonomy and dignity, justice, beneficence, and non-maleficence [15]. To ensure the reliability and validity of the results, different techniques were applied. These included developing a rapport with participants, creating a coding system, implementing peer review for themes and sub-themes, triangulation of data sources (including interview transcripts and field notes), and providing a thorough contextualised data description [16]. The specific names of the study public hospitals were not disclosed in the study to maintain anonymity. Therefore, the study hospitals remain unidentified to readers or any other individuals. The 12 interveiws participants (e.g., P12, P7, etc.), were also not identified by name, and the specific study site is known only to the researchers. Though, the ages of the 12 participants were mentioned in the participants’ quotes, given the large number of patients across the 21 public hospitals (from which the 3 study hospitals were selected), it is not possible for anyone to ascertain the age of each participant. The participants’ codes/IDs (e.g., P12, P7, etc.), we want to emphasize that these codes were assigned by the researchers solely for the purpose of this research. They are not related to any clinic or healthcare provider IDs used for medical follow-up. As a result, no individual, including the patients themselves, can identify any participant based on these codes.

## Results

### Socio-demographic characteristics of study participants

A total of 14 HIV-positive women with varying ages and educational levels participated in the in-depth interview. Their age range was from 20 to 59 years. Of the participants, 6 women were adhering and 6 women were non-adhering to their HAART medication.

Table 1 provides the demographic and background characteristics of the participants involved in the in-depth interview. The participants’ demographic and background characteristics suggest a diverse group with varying backgrounds and life experiences.

**Table 1.** Summary of demographic characteristics of HIV-positive women who participated in in-depth interviews.

| Characteristics (N=14) | Number | Total (%) |
| --- | --- | --- |
| **Age (yrs.)** |  |  |
| 20–29 | 3 | 25% |
| 30–39 | 5 | 41.7 % |
| 40–49 | 3 | 25% |
| 50–59 | 1 | 8.3% |
| **Adherence status** |  |  |
| Adherent to HAART | 6 | 50% |
| Non-adherent to HAART | 6 | 50% |
| **Educational level** |  |  |
| No education | 2 | 16.7% |
| Primary (1–8) | 5 | 41.7% |
| High school (9-12) | 4 | 33.3% |
| Tertiary (University) | 1 | 8.3% |
| **Marital Status** |  |  |
| Married | 4 | 33.3% |
| Divorced | 2 | 16.7% |
| Widowed | 3 | 25% |
| Single | 3 | 25% |
| **Employment status** |  |  |
| No work | 2 | 16.7% |
| Housewife | 4 | 33.3% |
| Labourer | 1 | 8.3% |
| Government | 0 | 0% |
| Private organisation | 5 | 41.7% |

### Themes, categories, and sub-categories

The experiences of HAART adherence among HIV-positive women were described in two themes. These themes include Barriers to HAART medication adherence and Facilitators of HAART medication adherence. Barriers to HAART medication adherence among HIV-positive women were grouped into five categories and fifteen sub-categories (Table 2). While facilitators of HAART medication adherence were grouped into five categories and seven sub-categories (Table 3).

**Table 2.**
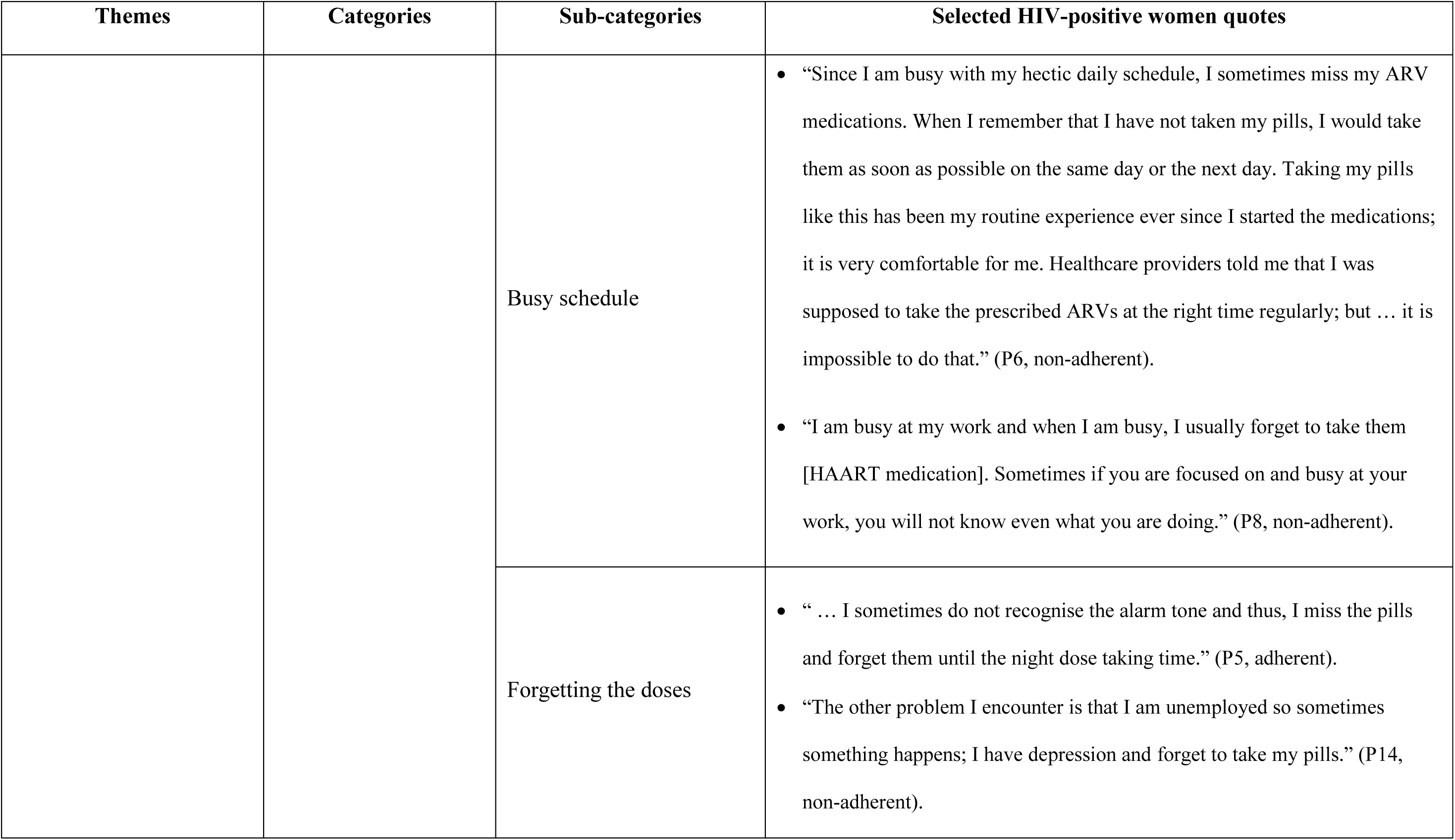
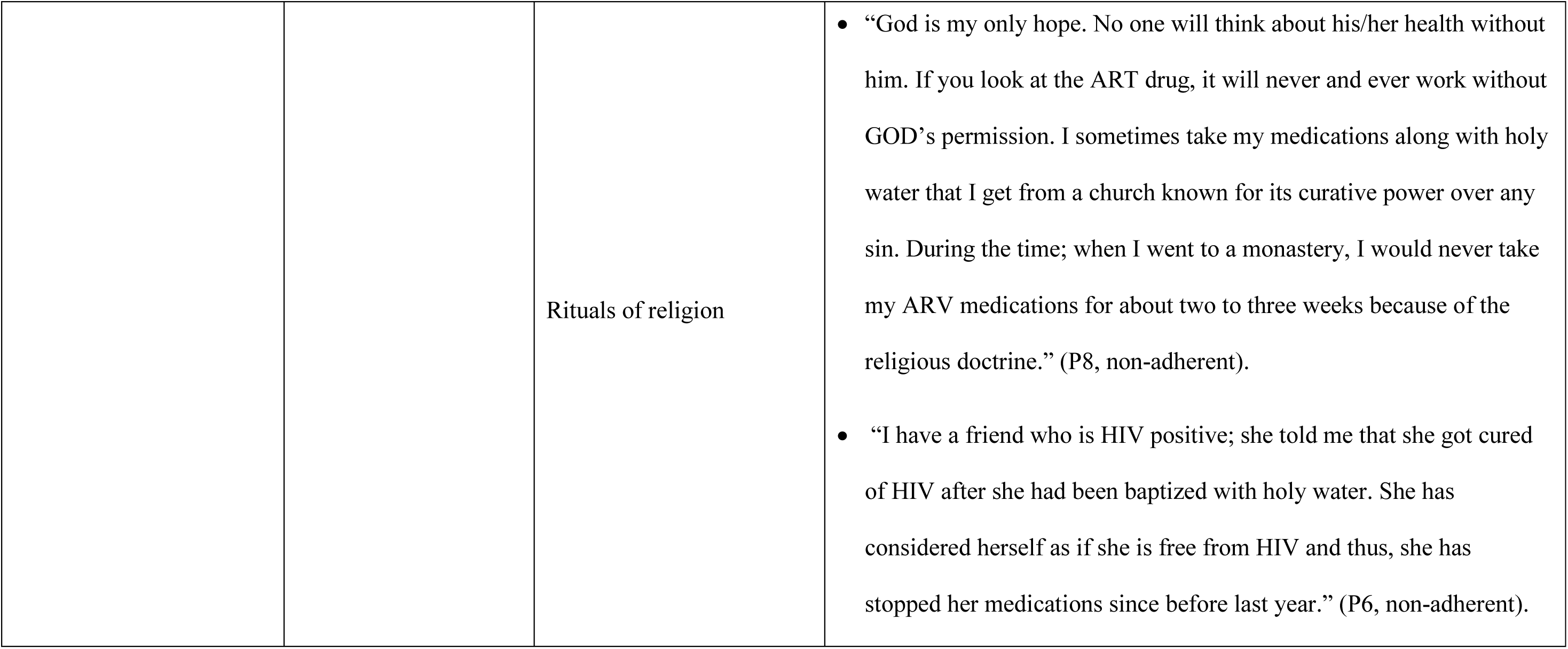
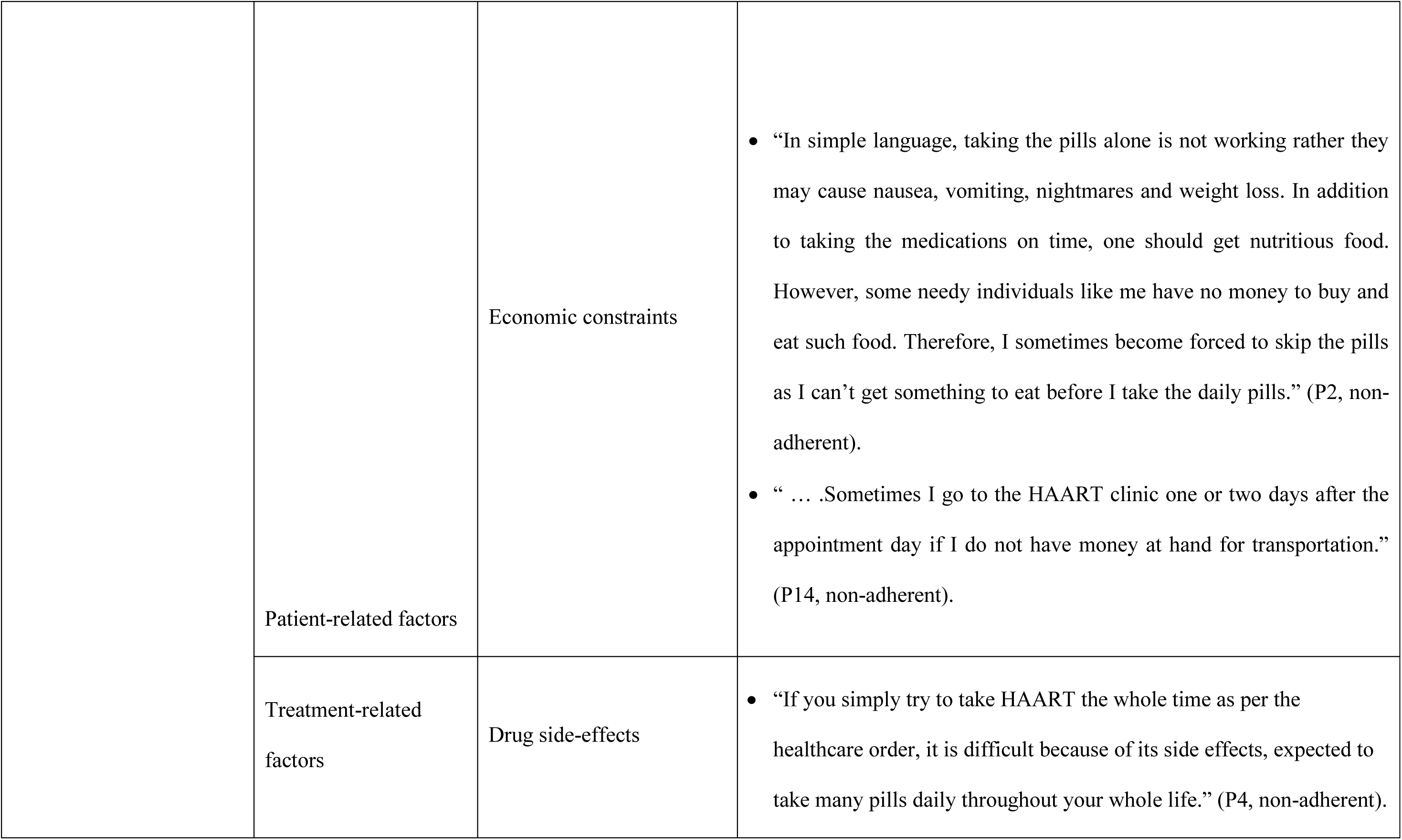
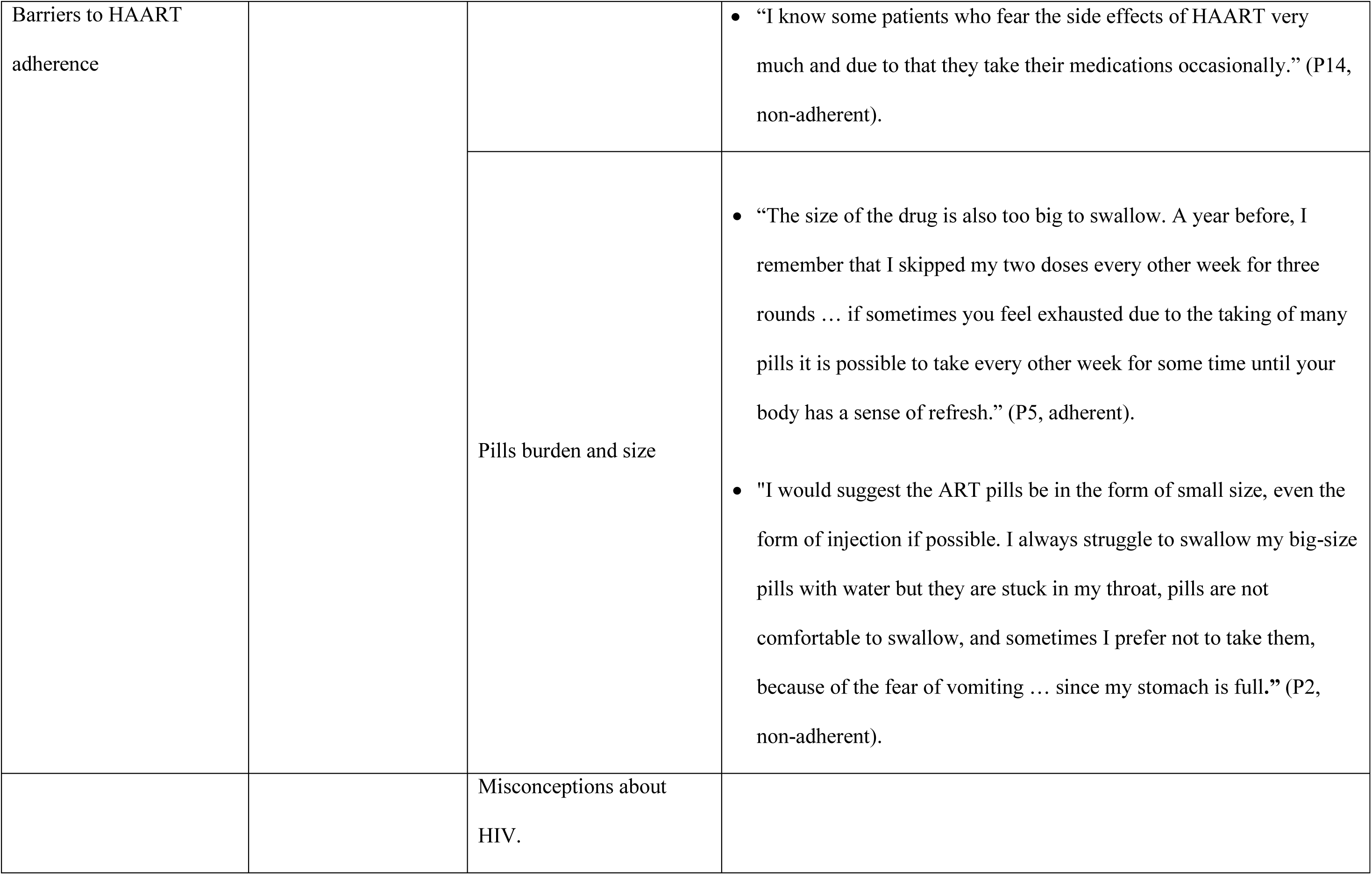
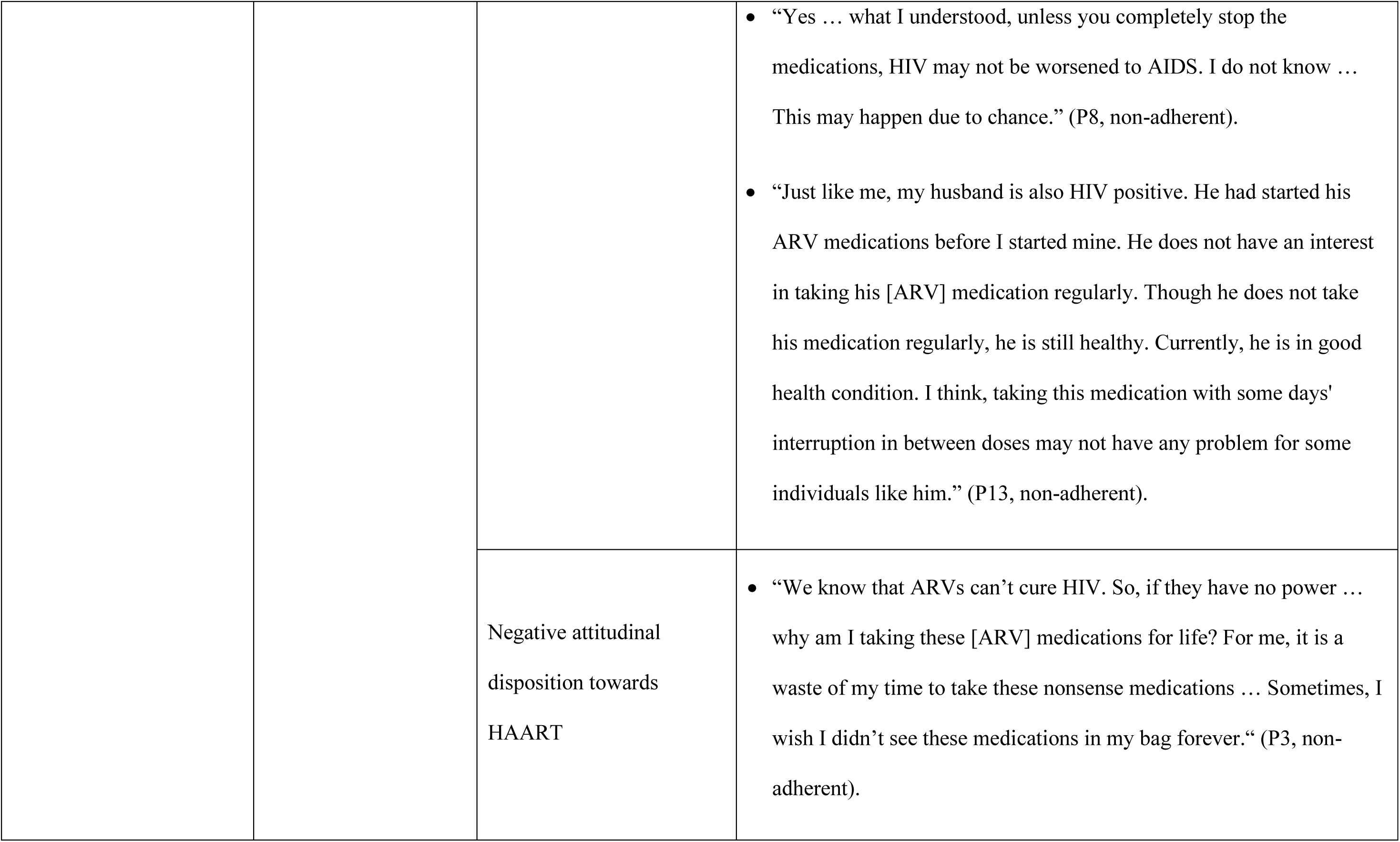
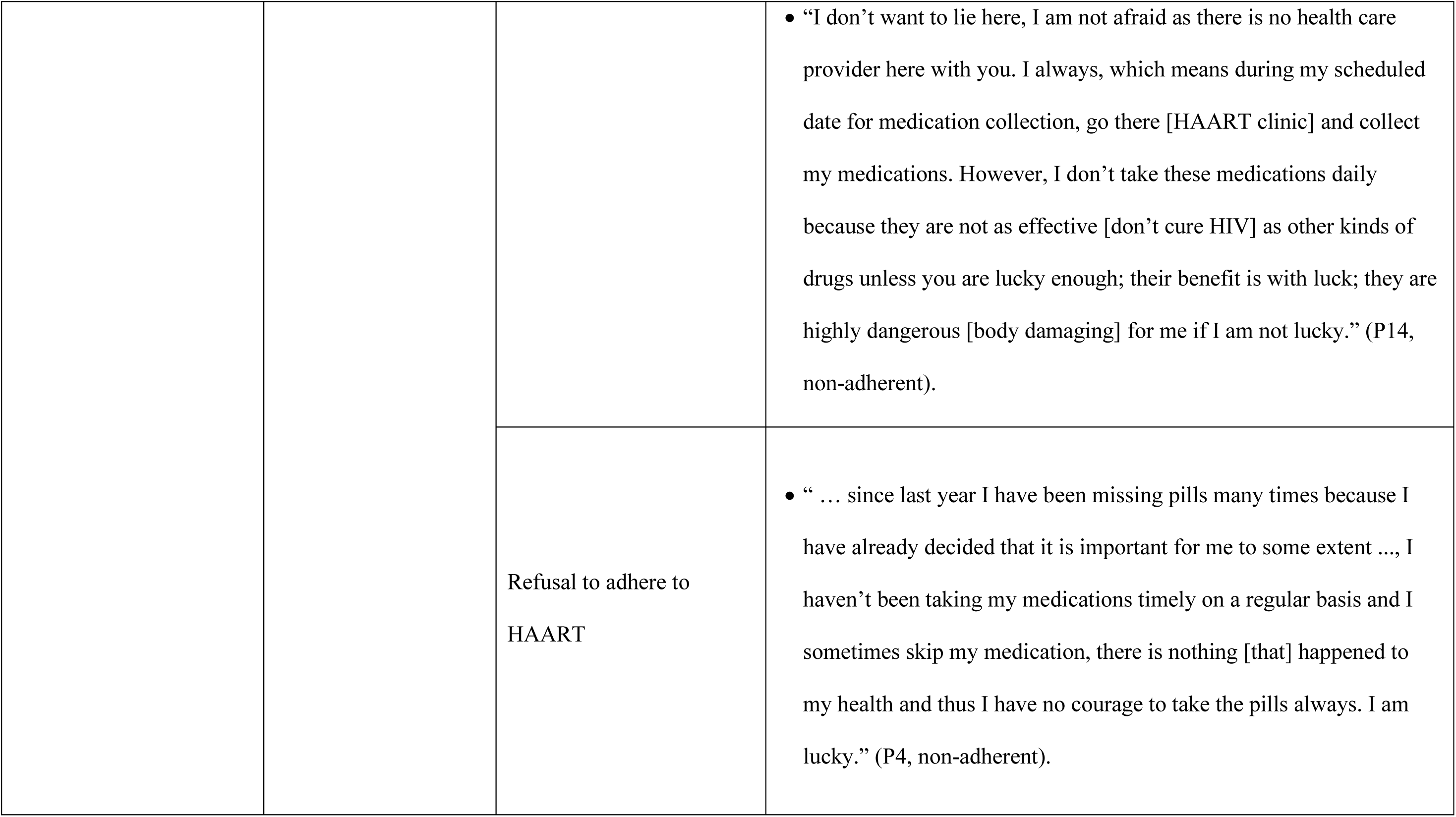
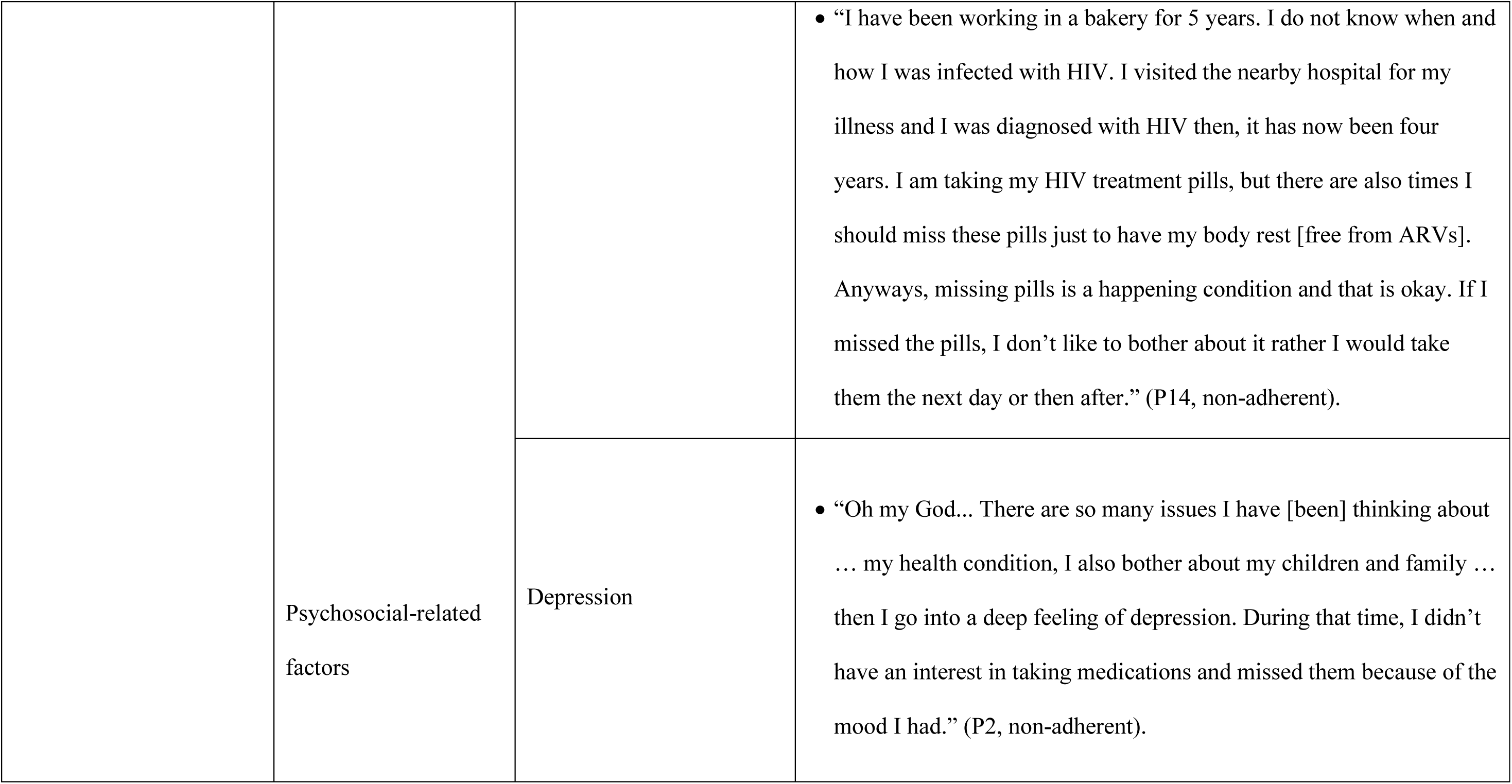
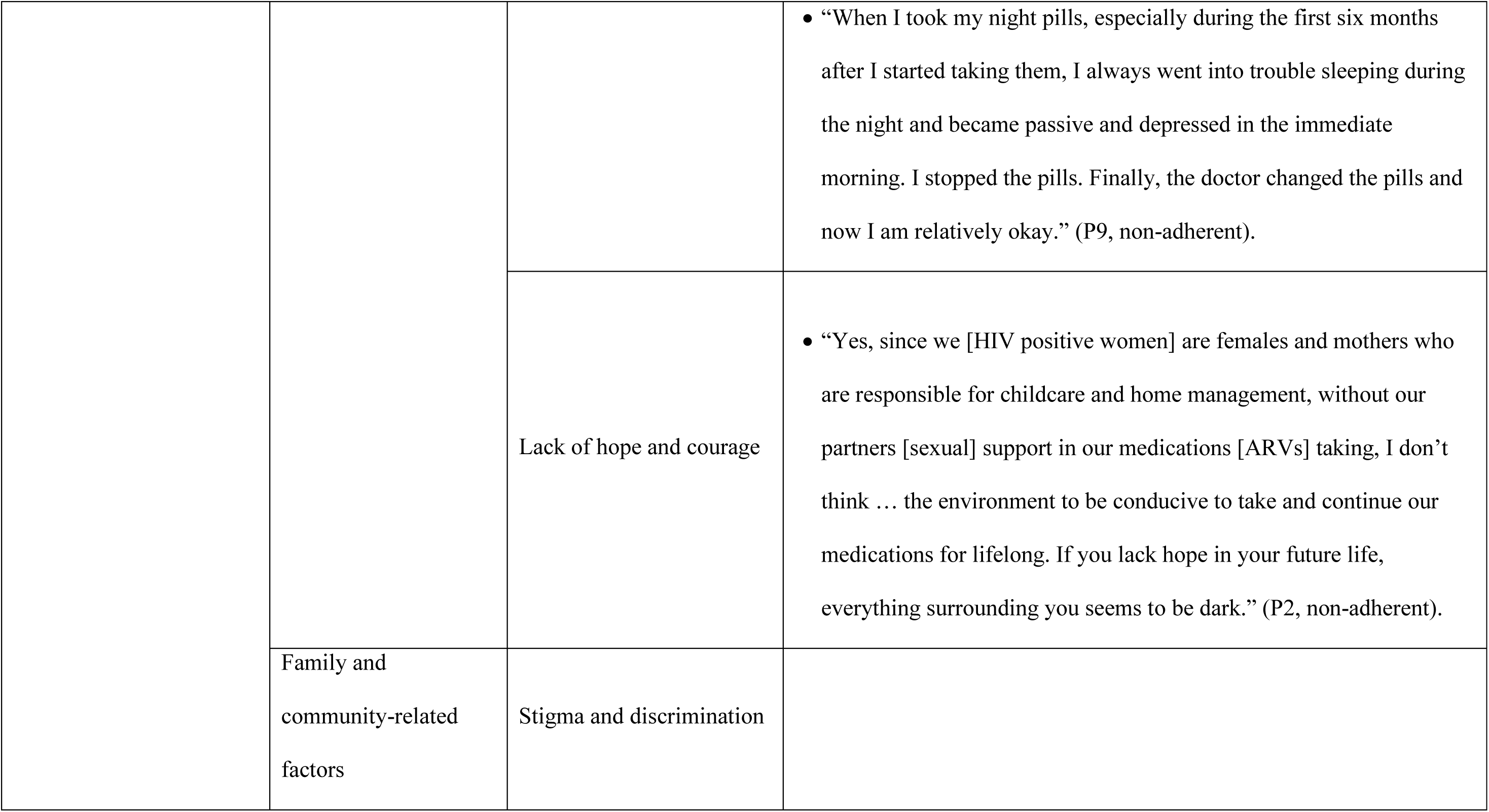
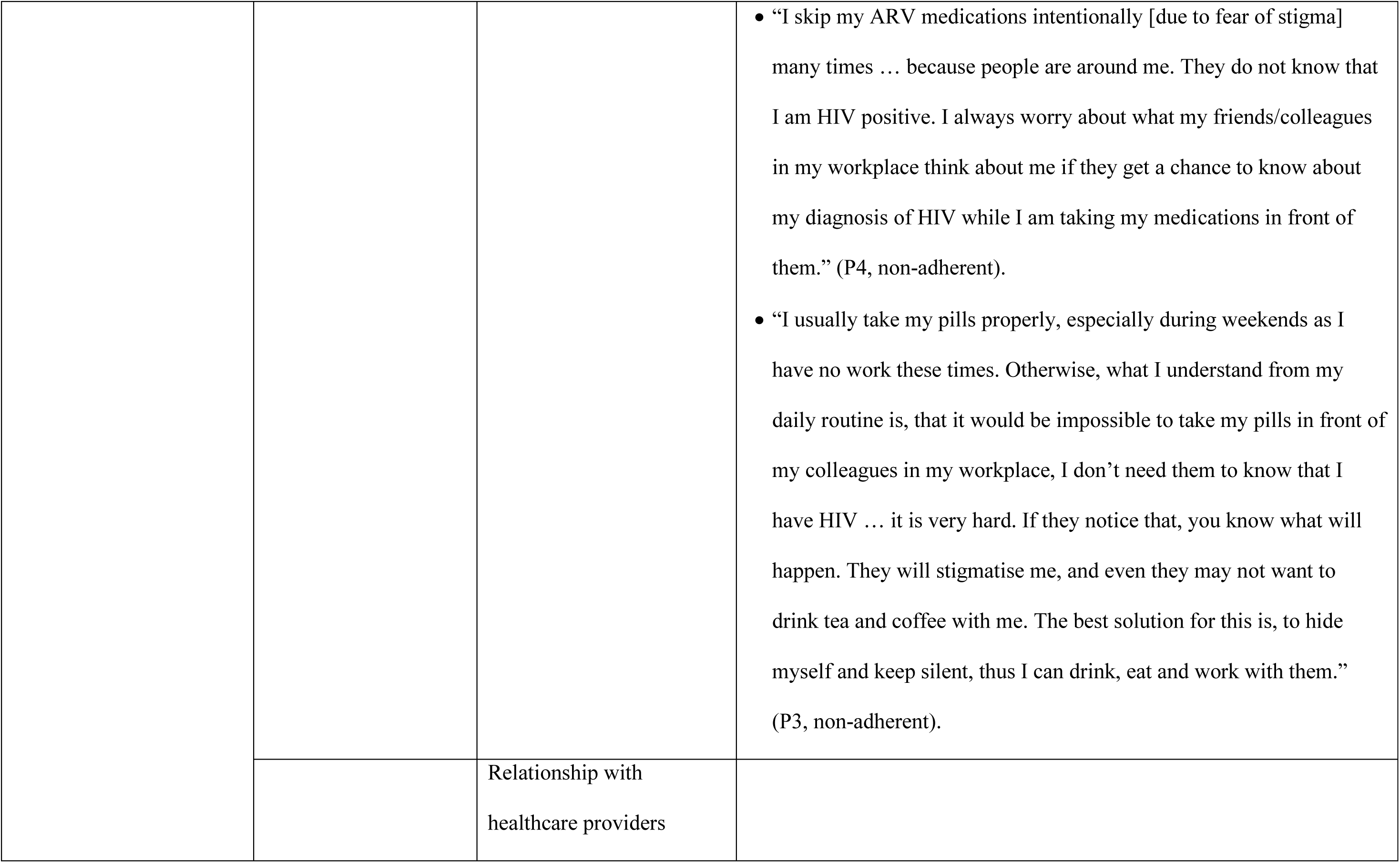
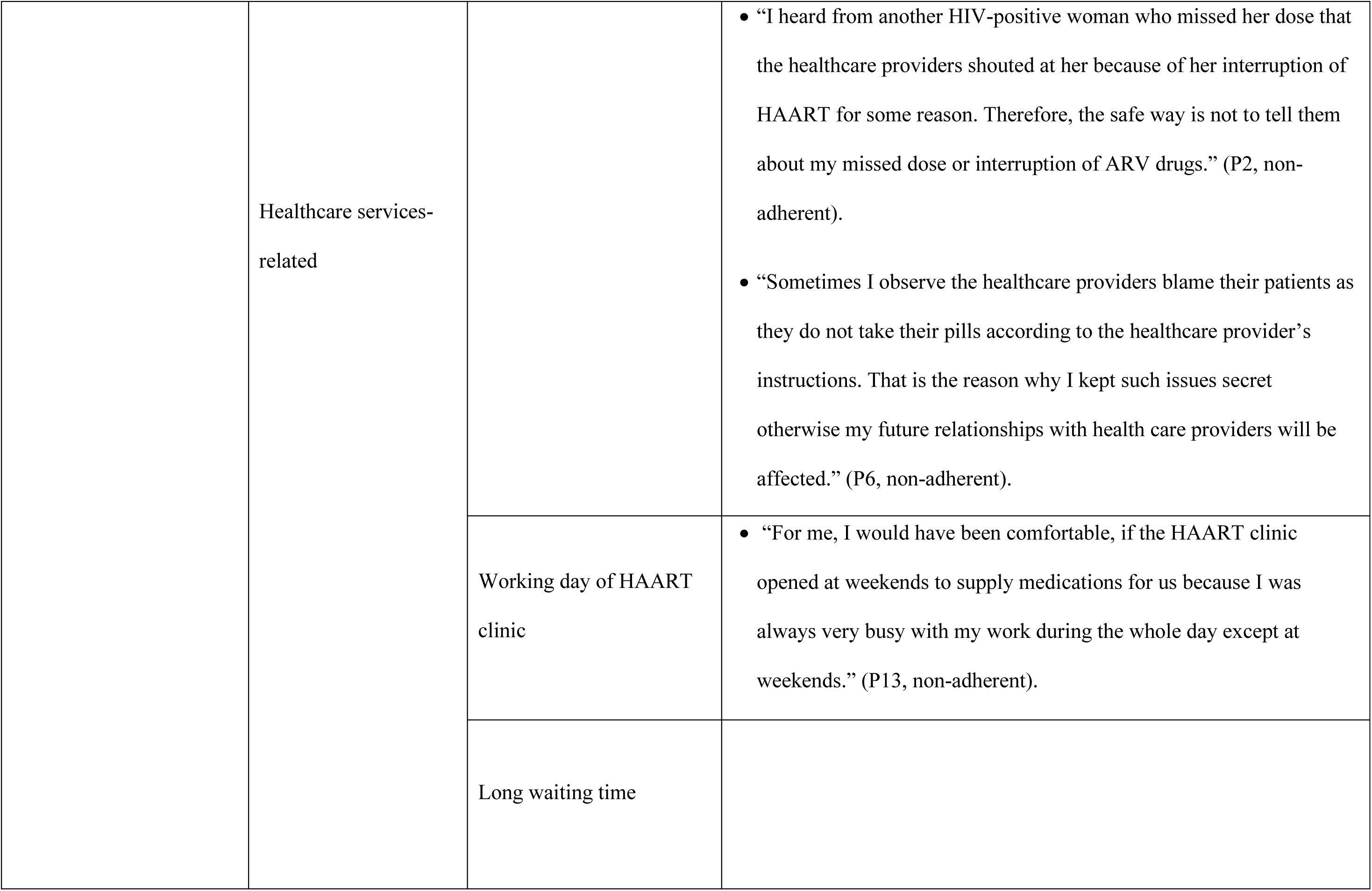
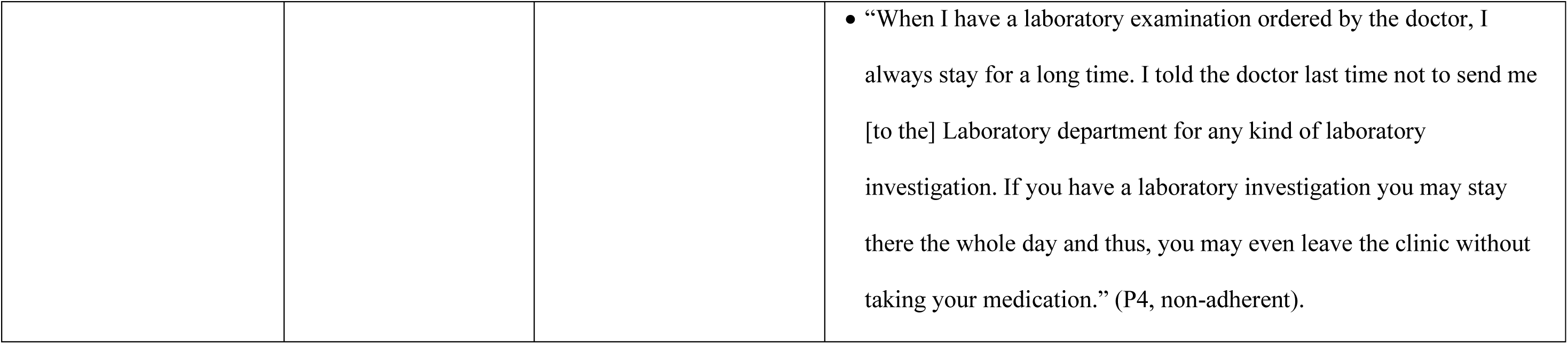
Barriers to HAART medication adherence among HIV-positive women in Southern Ethiopia.

**Table 3.**
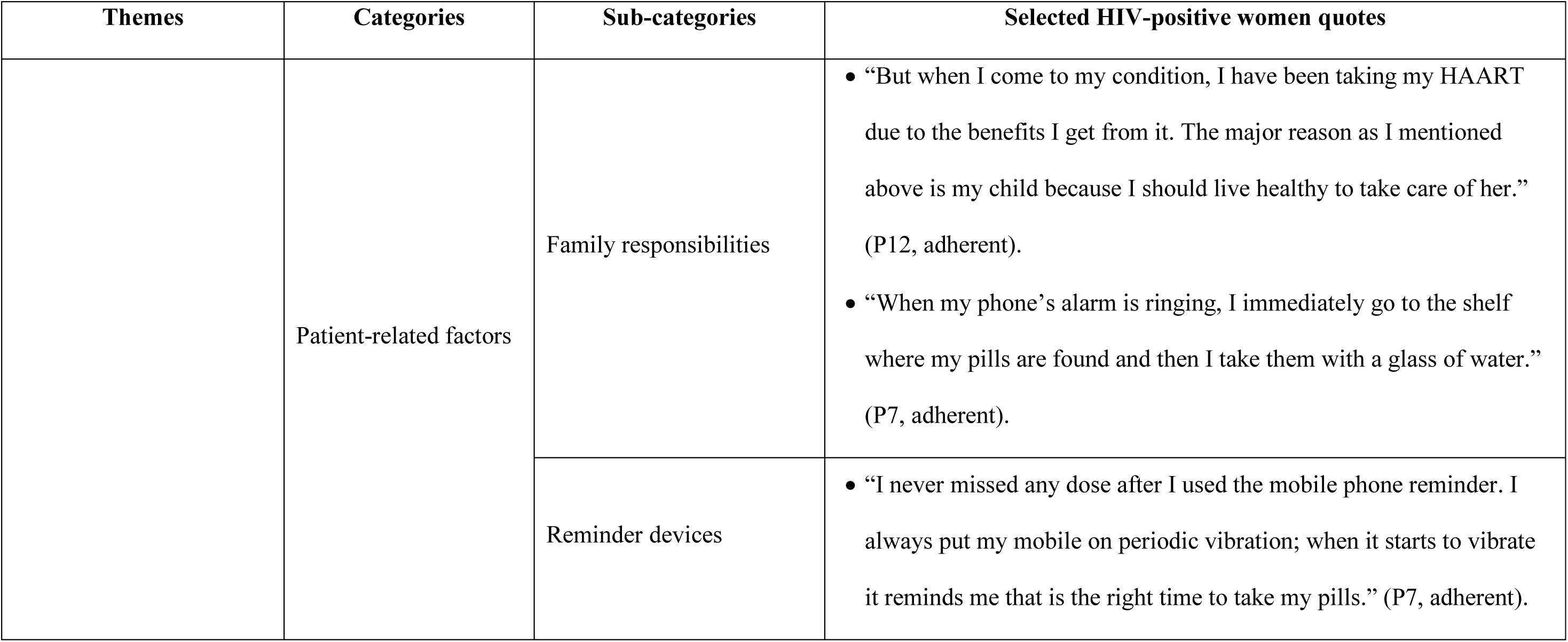
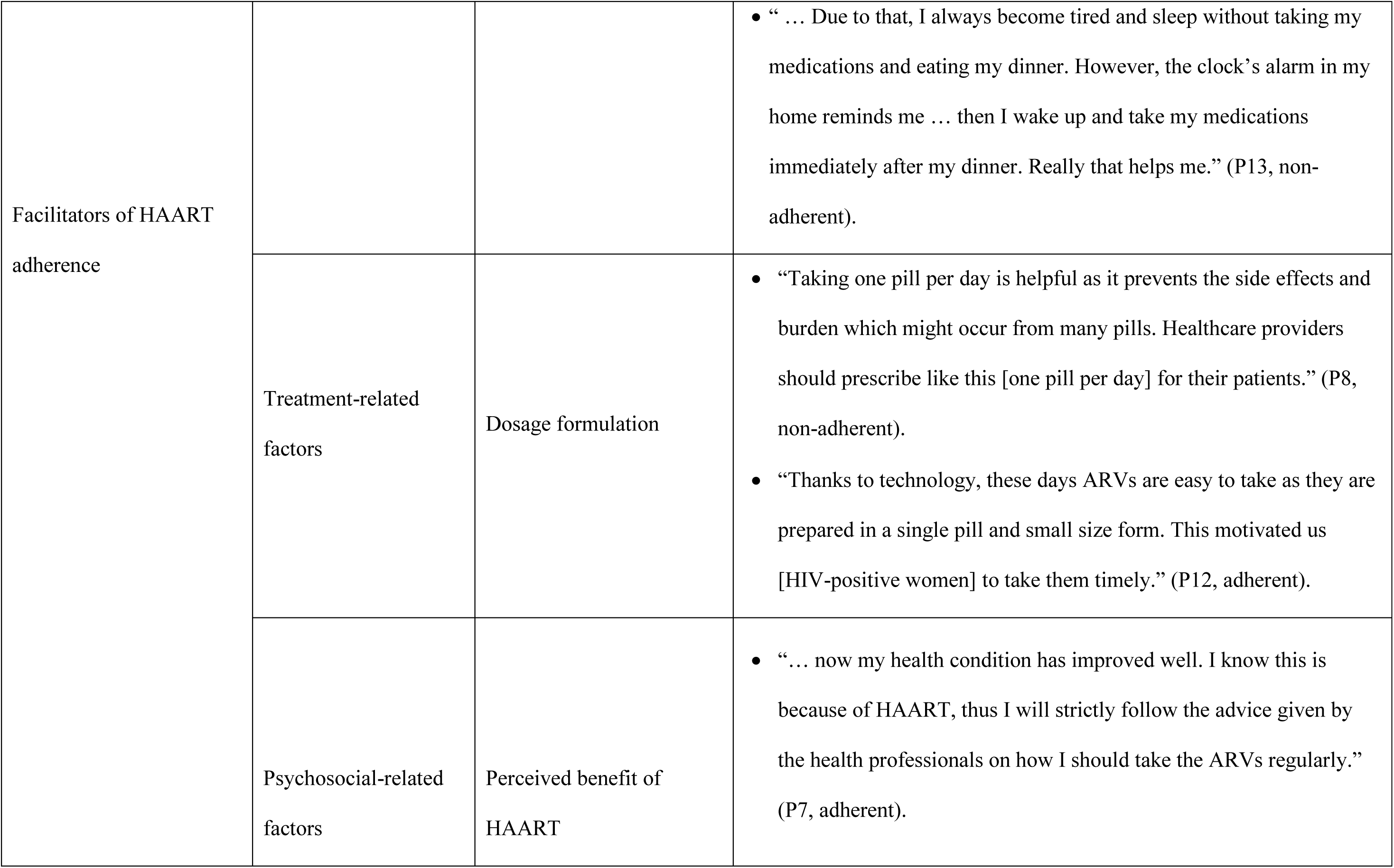
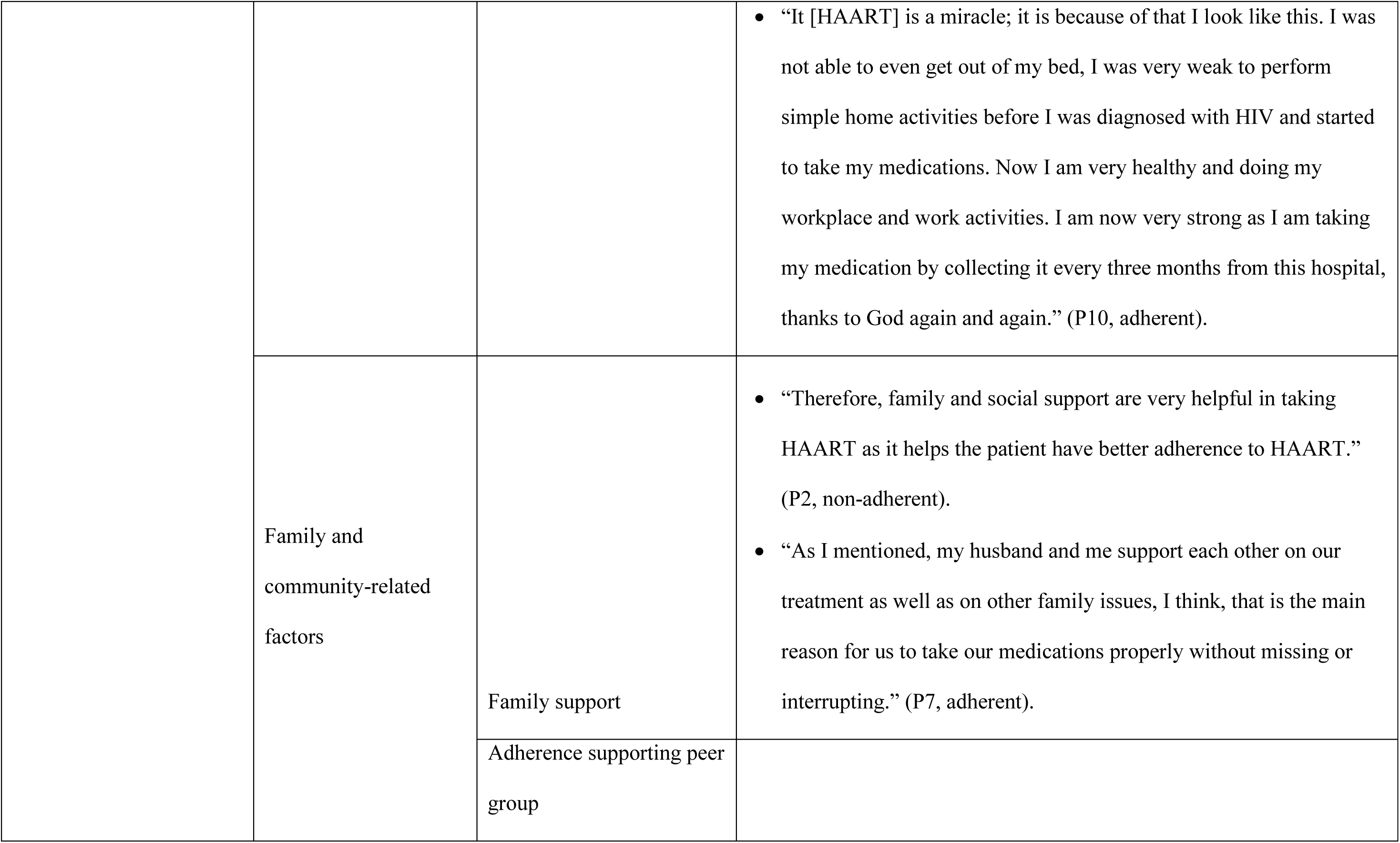
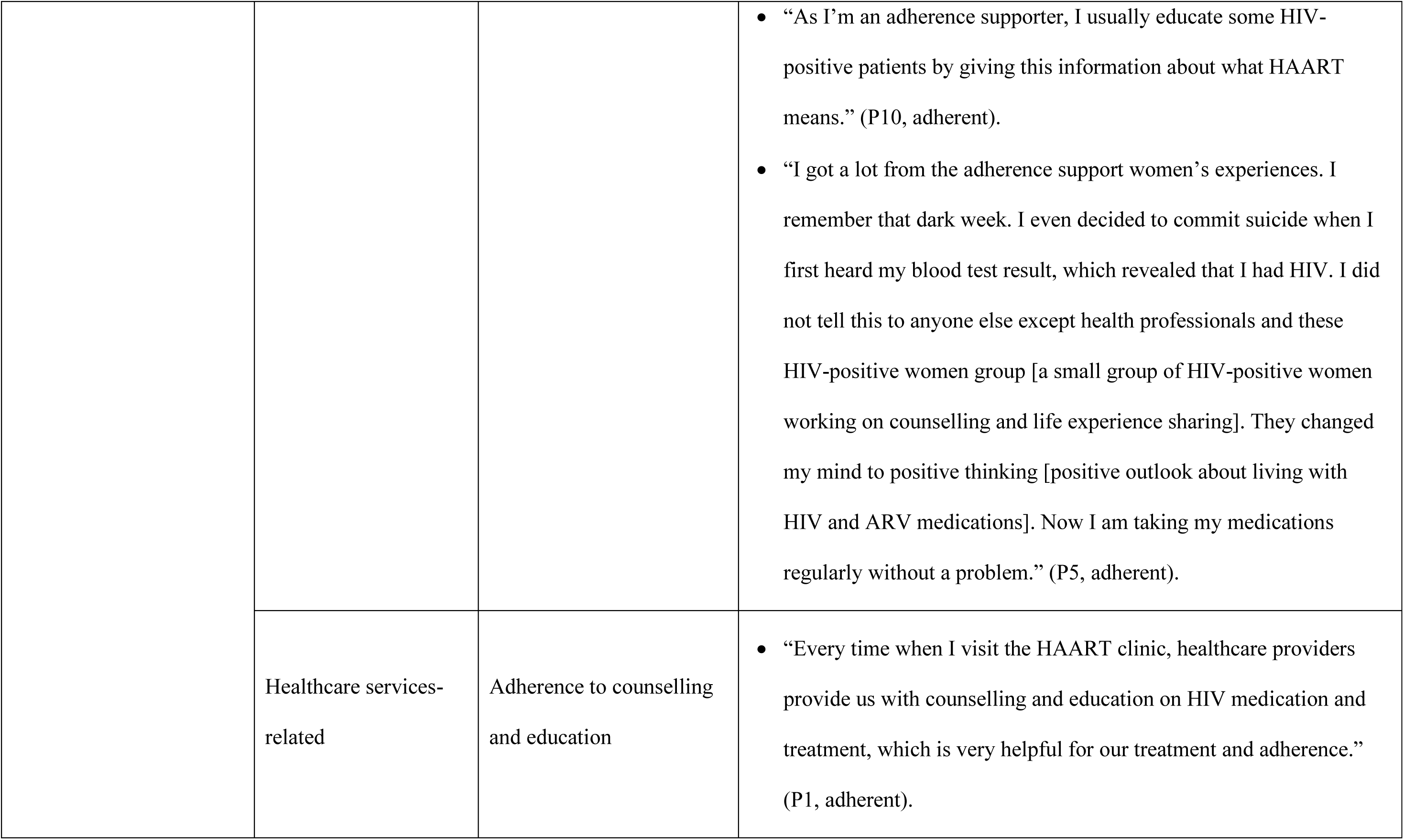
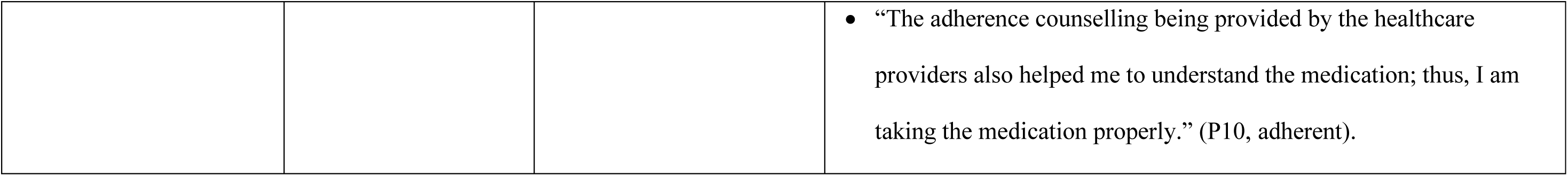
Facilitators to HAART medication adherence among HIV-positive women in Southern Ethiopia.

## Theme 1: Barriers to HAART Medication Adherence

### Category 1: Patient-related

#### Busy schedule

Participants (HIV-positive women taking HAART) reported some patient-related factors as barriers to their HAART medication adherence. These women highlighted their difficulties in keeping up with their HAART medication regimen due to their hectic schedules and employment obligations. Their hectic lifestyles frequently cause occasional missed dosages and difficulty adhering to the suggested right timing. During the IDIs, women discussed some barriers which impeded them from taking their HAART medication properly.

#### Forgetting the doses

Forgetting the HAART doses was among the issues reported by most participants in IDIs as a challenge for optimal HAART adherence. HIV-positive women may have a busy lifestyle that includes travelling somewhere away from their homes, engaging in different social affairs (weddings, funerals, social gatherings, and others), and personal reasons. Thus, these situations may create mental stress and insufficient time to remember their doses regularly, resulting in missed HAART doses.

One of the participants indicated that they occasionally forget to take their medication as directed because they get caught up in their everyday tasks or work.

#### Rituals of religion

Some participants of the IDIs highlighted how religious rituals, particularly fasting during Ramadan, or following Orthodox Church teachings, hindered their adherence to HAART medication. They believed that relying on their faith and religious practices would bring blessings and even potential cures for their health conditions. As a result, they chose to skip or delay taking their HAART medication during these religious periods. Despite their poor HAART adherence, some participants perceived themselves as healthy and attributed their well-being to the combination of religious rituals and medication.

#### Economic constraints

The study participants mentioned several financial barriers that prevented them from taking their HAART medication as prescribed by healthcare providers. Mentioning some symptoms like nausea, vomiting, nightmares, and weight loss as examples, they stressed the negative consequences of taking the medications without adequate nutrition. They frequently had to skip doses because their financial constraints made purchasing nutritious food impossible.

### Category 2: Treatment-related factors

#### Drug side-effects

Participants in the study expressed concerns about the side effects and organ damage that may result from using HAART medications. These worries and experiences exacerbate their difficulties in adhering to the HAART medication and the overall treatment plan. They indicated that missed doses would not worry them because their bodies would have a respite from the HAART medication.

#### Pills burden and pill size

Various reasons affect HIV-positive individuals’ adherence to their prescribed HAART medication regimen. The burden and size of pills are some of the factors that affect adherence to the lifelong treatment regimen for HIV/AIDS.

According to the participants’ views, taking HAART medication presented problems with their adherence due to the number and size of pills they had to take and their concerns about fatigue, discomfort, and swallowing big pills.

Some HIV-positive women who participated in the IDIs showed a misconception about the need for strict adherence to HAART medication. They think that periodic lapses in taking their HAART medication will not have a significant negative impact on their health or make their HIV situation worse. They attribute this belief to personal experiences or anecdotes of others who have seemingly maintained good health despite not strictly following the healthcare providers’ prescription for the HAART medication regimen.

### Category 3: Psychosocial-related factors

#### Negative attitudinal disposition towards HAART

In this qualitative study, some participants who took part in the IDIs expressed negative attitudes towards HAART medication, which hindered their HAART medication adherence. They expressed the negative attitude that HAART medication could not cure HIV and saw taking them as a waste of time, with potential long-term ineffectiveness and harm to their bodies.

#### Refusal to adhere to HAART

Refusal to adhere to HAART refers to the HIV-positive individual’s active decision not to adhere to the HAART medication as prescribed by healthcare providers. One major obstacle was the refusal to adhere, with some individuals choosing to miss their HIV treatment pills periodically to give their bodies a break from ARVs. Some held a mistaken belief that taking ARV medications daily without breaks could be harmful. They perceived no difference between daily and alternate-day intake of the pills and, as a result, decided to take their medications every other day. A barrier was a false sense of security, where certain participants believed they were lucky enough to evade negative consequences despite frequently skipping their medication. This belief resulted in a decreased drive to maintain consistent pill intake.

#### Depression

In the IDIs, depression has been identified as an obstacle preventing HIV-positive women from taking their HAART medication. Highly Active Antiretroviral Therapy (HAART) has been related to depression in some HIV-positive women, which has caused them to skip doses in order to prevent potential health issues:

#### Lack of hope and courage

Lack of hope and courage were found to be obstacles preventing study participants from taking their HAART medication. Some IDI participants expressed feelings of hopelessness, citing various reasons such as unsupportive partners, social pressure, the incurability of the disease, and fear of discrimination at work. Due to these factors and the gloomy outlook on their future, some HIV-positive women who participated in IDIs had become unmotivated to take their prescribed HAART medication consistently and even thought about stopping their treatment entirely.

### Category 4: Family and community-related factors

#### Stigma and discrimination

Many HIV-positive women indicated worries about taking their medications in front of coworkers at the workplace. They were concerned about possible negative consequences, including stigma, social exclusion, or rejection if others discovered their HIV status. Some HIV-positive women miss their medications on purpose due to this worry and apprehension.

### Category 5: Healthcare services-related

#### Relationship with healthcare providers

HIV-positive women looking for help and counselling for their health were facing challenges as a result of unpleasant encounters with healthcare providers. The challenges with HAART medication adherence have been exacerbated by such treatment at the HAART clinic because they feel discouraged and unsupported. The study participants reported several negative experiences with the healthcare providers.

#### Working days of HAART clinic (No service at weekends)

Due to the clinic’s operation hours conflicting with their working hours, many HIV-positive women, especially those who work during the week, find it challenging to pick up their ARV medications. Long absences from collecting medications as a result of this led to non-adherence.

#### Long waiting time

Participants often encounter long waiting times, especially during laboratory examinations or when there is a high influx of patients at the clinic. These extended waiting periods cause discomfort and inconvenience, making it difficult for them to access their medications and adhere to their HAART treatment prope

## Theme 2: Facilitators of HAART Medication Adherence

### Category 1: Patient-related

#### Family responsibilities

For many participants, the desire to be responsible for their children and ensure their well-being and education is a driver for maintaining a healthy life through proper medication adherence.

#### Reminder devices

In the IDIs, many participants reported that they use reminder devices. Particularly, a mobile phone alarm is a valuable reminder tool that could help HIV-positive women remember to take their HAART medication. HIV-positive women who participated in the IDIs underlined how useful these medication-reminder devices are for ensuring their regular and timely HAART medication intake.

### Category 2: Treatment-related factors

#### Dosage formulation

The dosage formulation of the HAART medication is crucial in enhancing patients’ compliance with their treatment regimen. The positive impacts of a simpler dosage formulation on participants’ adherence are evident from their feedback. Participants who had to take multiple drugs twice a day in the past often felt worn out and had problems adhering to the prescribed regimen. However, with the current single-pill dosage type, they take their medication very differently, which has increased adherence.

### Category 3: Psychosocial-related factors

#### Perceived benefit of HAART

Most participants were inspired to strictly follow their prescribed treatment plan by the benefits they observed from taking HAART medicine, particularly the notable health and general well-being improvement.

### Category 4: Family and community-related factors

#### Family support

Participants frequently stated that having family support has a good effect on their medication-taking habits and overall treatment experience. Participants with family support are encouraged to take their HAART medication as directed by their healthcare providers and are reminded to do so. Family members can actively ensure medication adherence, give prompt reminders, and foster a positive environment.

#### Adherence-supporting peer group

Participants highlighted the valuable support and guidance they receive from adherence-supporting peer groups, which positively influences their medication-taking behaviour. The adherence support women’s group significantly changed the participant’s perspective on coping with HIV and taking HAART medicine.

### Category 5: Healthcare services-related

#### Adherence counselling and education

Adherence counselling and education provided by the HAART clinic have encouraged participants to follow their HAART medication regimen. Continuous counselling and education have increased their awareness and consciousness regarding the importance of taking HAART medication properly. They emphasized that adherence counselling provided by healthcare providers could help them understand the importance of HAART medication and the consequences of poor medication adherence. It also helps them address any concern they have that needs medical attention.

## Discussion

In this study, HIV-positive women describe their experiences regarding their barriers and facilitators of HAART medication. These barriers include a busy schedule, forgetting the doses, rituals of religion, economic constraints, drug side-effects, pill burden and size, misconceptions about HIV, negative attitudinal disposition towards HAART, refusal to adhere to HAART, depression, lack of hope and courage, stigma and discrimination, relationship with healthcare providers, working day of HAART clinic, and long waiting times. Being busy with different activities linked to the roles of women was found to be one of the many challenges which affect the proper taking of medication among HIV-positive women. These findings of the IDIs are supported by the findings of two studies conducted in Nigeria. Being too busy was one of the reasons given by ARV users with poor adherence. Another study also identified being too busy as a significant factor influencing treatment adherence [17].

Forgetting to take HAART medication was also identified in the IDIs by participants as one of the reasons for poor medication adherence. Such problems associated with forgetting to take HAART medication in this study are consistent with findings from other studies [18, 19]. A study conducted in Northern Ethiopia found that forgetting and being away from home were prominent reasons identified for poor adherence among HIV-positive individuals receiving HAART [7]. Another barrier described by participants was spiritual activities. The spiritual activities including fasting and the use of holy water, are also mentioned in another study from Northwest Ethiopia as challenges to HAART adherence among HIV-positive individuals [20]. A study conducted in London found that some HIV-positive individuals on HAART believed their faith in God might cure HIV/AIDS [21]. Economic constraints were among the barriers to HAART medication raised by the participants in this study. This finding is consistent with another study conducted in Uganda that the Lack of money for transport was reported as a challenge to HAART adherence [22]. Furthermore, financial challenges were one of the major barriers to HAART medication adherence among HIV-positive individuals [23].

Drug side effects appeared to be one of the major barriers to HAART medication raised by the participants in this study. This is also mentioned in the literature [24]. Those women who had developed side effects while on treatment had poor adherence to their HAART medication [25]. Moreover, other studies conducted in Ghana have revealed that medication side effects harm HAART adherence [26, 27]. The current study also explored pill burden and pill size as among the common challenges of HIV-positive women in taking their HAART medication. This challenge affects the day-to-day activities, especially the daily taking of ARV pills, and thus affects their adherence to HAART. If individuals diagnosed with HIV have some inconvenience with their HAART medication regimen, the likelihood of taking their medication at the right time and frequency will be reduced, which results in poor [21] adherence to HAART medication [28]. A similar study conducted in Dessie also indicates that HIV-positive individuals who took medications other than HAART medication were less likely to adhere to their HAART medication compared to those who did not take any non-HAART medications [29]. Some HIV-positive women who participated in this study showed a misconception about the need for strict adherence to HAART medication. Negative attitudinal dispositions towards HAART were also reported by participants as a barrier to their HAART. The study showed that participants think that periodic lapses in taking their HAART medication will not have a significant negative impact on their health or make their HIV situation worse. It is revealed that misconceptions can lead to HIV-positive individuals delaying or avoiding treatment of HIV with HAART medication, which can have negative consequences for their health and well-being. Negative perceptions or beliefs of HIV-positive individuals of the personal need for HAART medication in HIV-positive individuals cause poorer adherence to their HAART medication regimen [30]. HIV-positive individuals’ negative perception toward HAART was identified in this study as one of the multiple factors which hinder HAART medication adherence. This factor has also been reported in other similar studies [31]. Moreover, the refusal to adhere was explored as a barrier to HAART adherence in this study. Some individuals choose to miss their HIV treatment pills periodically to give their bodies a break from ARVs and this results in poor adherence. This study’s findings support the findings from other studies [32, 33]. On the other hand, depression, lack of hope and courage and stigma and discrimination were raised as common barriers to HAART medication by the participants. Depression is a psychiatric disorder that manifests clinically as low mood, loss of enjoyment, decreased motivation and energy, guilt or low self-esteem, disturbed sleep or appetite, suicidal thoughts, and trouble concentrating [34]. A combination of factors, including the nature of the disease, medication side effects, social stigma and discrimination impacts their adherence to HAART negatively [35, 36]. Similarly, a study conducted in Eastern Ethiopia also reported depression as one of the factors that affect ART adherence [37]. When partner support is lacking, several challenges may arise that could affect treatment adherence, like the lack of emotional support and encouragement [38]. HIV stigma is a common problem and frequently reported barrier to HAART medication adherence among HIV-positive individuals. A study done in Kenya revealed that HIV-related stigma was found to be the major challenge for HIV-positive individuals, preventing them from taking their HAART medication properly as prescribed by healthcare providers [39]. The influence of stigma and discrimination on HAART adherence is also shared by other studies [40]. In this study, participants reported poor relationships with the healthcare providers and due to this their medication-taking practices have been affected. These findings are in support of studies also documenting that poor relationships between healthcare providers and patients can negatively impact drug adherence [41]. The absence of services over weekends and long waiting times at HAART clinics were mentioned as barriers to HAART medication among participants. A qualitative study indicated that some patients do not have flexible work schedules that allow them to pick their medication up during clinic weekday [42]. Long waiting times at HAART clinics are a significant factor promoting poor adherence to HAART medication, according to studies in Ghana and in South Africa [43, 44].

Participants described facilitators of HAART adherence as family responsibilities, reminder devices, dosage formulation, perceived benefit of HAART, family support, adherence supporting peer groups, and adherence to counselling/education. A qualitative study conducted in Zimbabwe indicated that HAART helped HIV-positive women carry out their responsibilities as mothers in caring for their children and family members [45]. In this study, participants underlined how useful these medication reminders were to ensure their regular and timely HAART medication intake. The importance of reminder devices in medication-taking practices was revealed in different studies [46, 47]. Likewise, the current study findings indicated that a mobile phone alarm is a valuable reminder tool that could help HIV-positive women remember to take their HAART medication.

In this study, participants who had to take multiple drugs twice a day in the past often felt worn out and had problems adhering to the prescribed regimen. However, with the current single-pill dosage type, they take their medication very differently, which has increased adherence. Single-tablet regimens encourage treatment adherence since they are easier to use and have reduced toxicities [48]. On the other hand, many participants reported improvements in both their physical and overall health conditions as a result of HAART. The findings of this study on the benefit of HAART are similar to other studies [49]. Researchers found a positive association between family support and adherence to HAART medication [50]. A qualitative study in South Africa showed that patient education and counselling in the HAART clinic facilitate treatment adherence [51]. In this study, it is reported that adherence counselling and education provided by the HAART clinic have encouraged participants to follow their HAART medication regimen.

## Limitations

The study was restricted to and conducted in only three urban hospitals in one region of Ethiopia. Therefore, the findings cannot be generalised to other regions or the whole country. Another limitation of the study was the difficulty in avoiding the possibility of social desirability bias. This was during the face-to-face in-depth interviews, particularly from HIV-positive women who came to HAART clinics to collect their HAART medication, which might increase the chance of interviewees answering questions with more desirable responses instead of actual answers. Despite these limitations, we used face-to-face in-depth interviews with participants provided a rich and personal understanding of their experiences, allowing for a deeper exploration of the factors influencing adherence to HAART medication.

## Conclusions

Adherence to HAART medication is a major challenge among HIV-positive women in Southern Ethiopia. In exploring the HIV-positive women’s own experiences and needs an understanding of the barriers and facilitators of adherence to HAART medication can be reached. Busy schedule, forgetting the doses, rituals of religion, economic constraints, drug side-effects, pills burden and size, misconceptions about HIV, negative attitudinal disposition towards HAART, refusal to adhere to HAART, depression, lack of hope and courage, stigma and discrimination, relationship with healthcare providers, a working day of HAART clinic, and long waiting time were identified as barriers to HAART adherence. While, family responsibilities, reminder devices, dosage formulation, perceived benefit of HAART, family support, adherence supporting peer groups, and adherence to counselling/education were identified as facilitators of HAART adherence. Therefore, understanding factors which affect adherence can help to develop tailored interventions to increase adherence and improve the health outcomes of HIV-positive women.

## Data availability statement

The raw data supporting the conclusion of this article will be made available by the authors, without undue reservation.

## Acknowledgements

We would like to express our gratitude to all the three sampled public hospitals’ healthcare providers, all the participants and their families, and all of our colleagues who contributed to this study.

## Author Contributions

**Conceptualization:** Alemayehu Abebe Demissie.

**Data curation:** Alemayehu Abebe Demissie.

**Formal analysis:** Alemayehu Abebe Demissie and Elsie Janse van Rensburg.

**Investigation:** Alemayehu Abebe Demissie.

**Methodology:** Alemayehu Abebe Demissie and Elsie Janse van Rensburg.

**Supervision:** Elsie Janse van Rensburg.

**Writing – original draft:** Alemayehu Abebe Demissie.

**Writing – review & editing:** Elsie Janse van Rensburg.

